# Cohort Profile: Investigation into Biomarkers to Predict Preterm Birth (INSIGHT) -- a Prospective Pregnancy Cohort Focused on Preterm Birth in the United Kingdom

**DOI:** 10.64898/2026.04.08.26350031

**Authors:** Raphaella Jackson, Carlotta Valensin, Evonne Chin-Smith, Natalie Suff, Andrew H. Shennan, Natasha L. Hezelgrave, Rachel M. Tribe

**Affiliations:** King’s College London, Department of Women and Children’s Health, London, United Kingdom; Precision Genomics and Translational Omics Lab, Sidra Medicine, Doha, Qatar; Centre for Fetal Care, Queen Charlotte’s and Chelsea Hospital, Imperial College Healthcare NHS Trust, London, United Kingdom

## Abstract

**Purpose:** Spontaneous preterm birth (sPTB), particularly early preterm birth and mid-trimester loss, remains poorly understood and difficult to predict. The INSIGHT cohort was established to create a deeply phenotyped, longitudinal pregnancy dataset integrating clinical data and biological sampling to investigate the mechanisms of cervical shortening and sPTB, with a focus on linking innate immune responses, the vaginal microbiome, and host biology to identify early biomarkers of risk.

**Participants:** 2272 pregnant women (8^+0^–28^+0^ weeks’ gestation) were enrolled as high or low risk of preterm birth based on obstetric history, cervical length, cervical procedures, multiple pregnancy, or Müllerian anomalies. Serial clinical data and biological samples, including cervicovaginal specimens and blood, were collected throughout pregnancy.

**Findings to date:** The cohort has generated comprehensive multi-omic data, including transcriptomic, microbiome, metabolomic, proteomic, and immune profiling. Key findings demonstrate that maternal plasma cfRNA can predict early sPTB months before clinical presentation, and that integration of cervicovaginal microbiota, metabolites, and host immune markers improves risk prediction and provides mechanistic insight into inflammatory pathways leading to sPTB.

**Future plans:** Recruitment concluded in 2023, with final visits occurring in 2024. Ongoing analyses focus on refining predictive models, defining biological subtypes of preterm birth, and translating integrated biomarker panels into clinically scalable risk stratification tools.

**STRENGTHS AND LIMITATIONS OF THIS STUDY:**

- Large, prospective longitudinal cohort (Strength): Ten years of recruitment with repeat sampling enabled detailed study of biological pathways leading to sPTB.
- Broad risk spectrum with clear definitions (Strength): Inclusion of both high and low-risk women using pre-specified clinical criteria supported robust comparative analyses and biomarker discovery.
- Multicentre NHS recruitment (Strength): Inclusion of several sites, particularly the diverse Lambeth population at St Thomas’, enhanced population diversity and external validity.
- Hospital-based, high-risk enrichment (Limitation/Strength): Recruitment from specialist preterm birth clinics and secondary/tertiary care may limit generalisability to lower-risk or primary care populations. However, it did ensure many preterm birth events were captured prospectively in this study.
- Incomplete follow-up and limited late sampling (Limitation): Attrition and sampling only up to a prespecified gestation (defined by standard clinical pathway) reduced full pregnancy coverage of longitudinal data.

## INTRODUCTION

Preterm birth, defined as birth before 37 weeks of gestation, is one of the most complex and pressing challenges in modern obstetric medicine. It is a leading cause of neonatal morbidity and mortality worldwide, with substantial long-term consequences for affected children and significant economic and health system burdens (1–4). Despite decades of research, the mechanisms of preterm birth remain poorly understood due to the complex and heterogeneous aetiologies involved (5). Current interventions and predictive tools remain limited in accuracy and scope, highlighting the need for further developments to effectively manage this global health concern (6,7).

The INSIGHT cohort study was established to address this need by creating a deeply phenotyped cohort with comprehensive longitudinal data and biological sampling to investigate the mechanisms of spontaneous preterm birth in women at higher risk of early delivery due to obstetric and medical history. This risk group has higher rates of preterm birth, particularly mid-trimester loss and early preterm birth (8–10) and results in some of the most severe consequences for affected mothers and children. Mechanisms driving early sPTB are multifactorial but are most likely to be associated with a complex interaction of maternal and/or fetal inflammatory responses which culminate to the final common pathway of progressive cervical shortening and myometrial contractions (11). The principal aim of the study was to link innate immune function and host response with vaginal microbiome populations and gene polymorphisms. To achieve this, women at low and high risk of preterm birth were recruited to provide clinical data and a range of biological samples were collected from participants at various timepoints throughout their pregnancy including: cervicovaginal cells/fluid/mucus and blood.

Here, we provide a comprehensive overview of the INSIGHT cohort study and current findings. We describe the population and core outcomes, alongside evaluating the impact of known risk factors. We outline the range of biological samples collected and research output to date.

## COHORT DESCRIPTION

### 5. Study Overview

The INSIGHT study was a prospective cohort study which aimed to determine the biological mechanisms underlying cervical shortening and spontaneous preterm birth, along with identifying novel biomarkers which could identify women at high risk for preterm birth early in their pregnancy. The study was primarily conducted at Guy’s and St Thomas’ NHS Foundation Hospital Trust (GSTT) in London with additional recruitment at St. Mary’s Hospital (Manchester NHS Foundation Trust), St Mary’s Hospital (NHS University Hospitals Dorset NHS Foundation Trust), and West Middlesex University Hospital (Chelsea and Westminster Hospital NHS Foundation Trust). The study was active for years between 2013 and 2024 (recruitment finished in 2023).

### 6. Recruitment and Eligibility

INSIGHT recruited pregnant women between 8^+0^ and 28^+0^ weeks’ gestation, classified into two groups according to their risk of preterm birth. High-risk status was defined by the presence of one or more of the following criteria:

- Previous preterm birth or mid-trimester loss between 16^+0^ and 37^+6^ weeks’ gestation
- Previous preterm premature rupture of the fetal membranes (PPROM) at ≤ 37+6 weeks’ gestation
- Short cervical length (≤ 25 mm) on ultrasound between 18^+0^ and 24^+0^ weeks’ gestation
- Previous cervical procedure to treat abnormal smears (large loop excision, laser conisation, cold knife conisation, or radical diathermy)
- Multiple pregnancy
- Known Müllerian Abnormalities

Low-risk participants were those who met none of the above criteria. Women were excluded if there were known significant congenital, structural, or chromosomal fetal abnormalities. Potential high-risk participants were identified from referral letters to, or attendance at, preterm birth clinics, while potential low-risk participants were identified through attendance at antenatal clinics, early pregnancy units, or antenatal day units. In both cases, prospective participants were informed of the study and offered the opportunity to take part. Figure 1 shows the flow of participants from consent through to enrolment, together with total recruitment by site.

**Figure 1:**
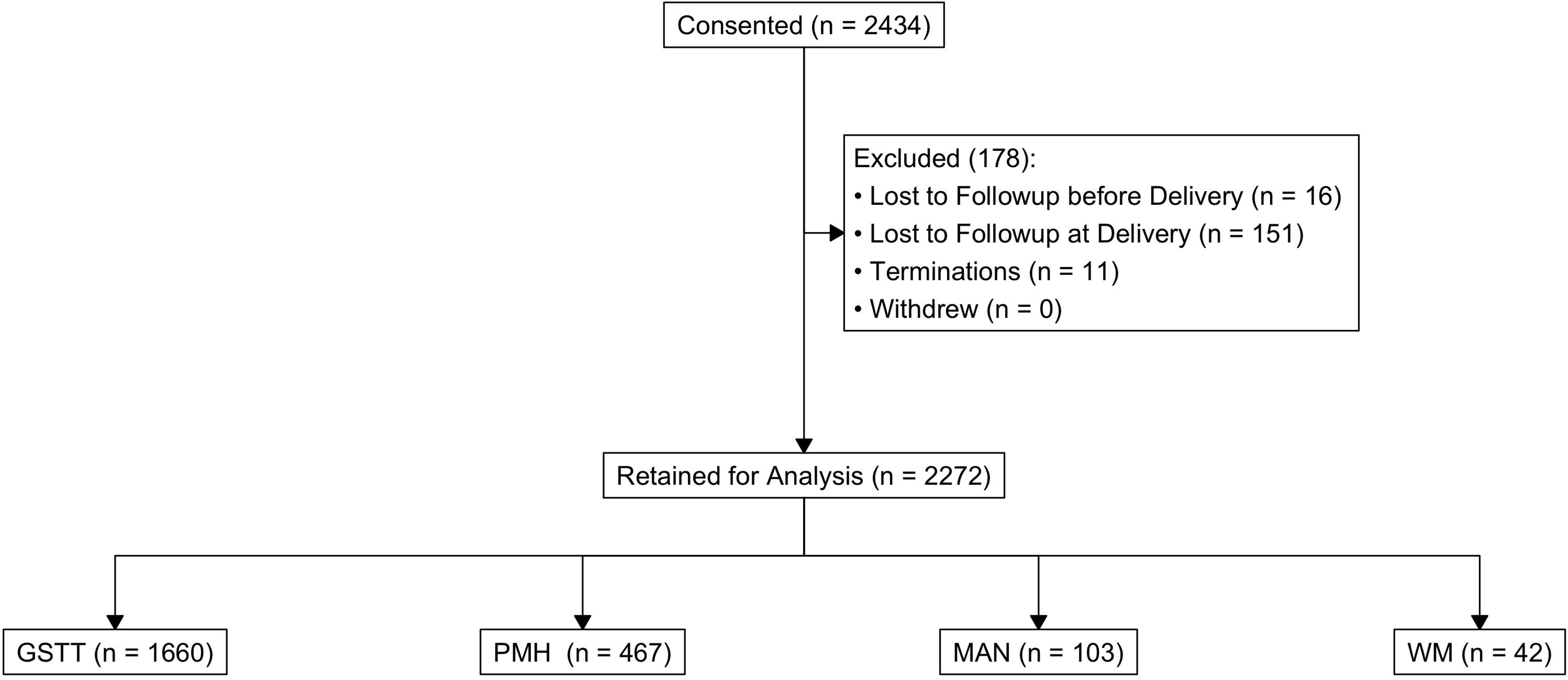
A consort plot showing the flow of participants from consent to analysis, including a breakdown of participants per site of recruitment. Abbreviations: GSTT = Guy’s and St Thomas’ NHS Foundation Hospital Trust (GSTT) London, MAN = St. Mary’s Hospital (Manchester NHS Foundation Trust), PMH = St Mary’s Hospital (NHS University Hospitals Dorset NHS Foundation Trust), WM = West Middlesex University Hospital (Chelsea and Westminster Hospital NHS Foundation Trust).

### 7. Participant Characteristics

#### a. Characteristics at Study Intake

A broad set of baseline variables spanning sociodemographic background, clinical context at enrolment, and index pregnancy details were collected. These included age at registration, ethnicity, and residential lower super output area, a small-area UK geographic unit that enables linkage to area-level deprivation indices and other geographically referenced data. Routine anthropometric measures (height, weight, and derived BMI) were also recorded given their established associations with obstetric complications, including preterm birth. A selection of these baseline characteristics are summarised in Table 1.

**Table 1:**
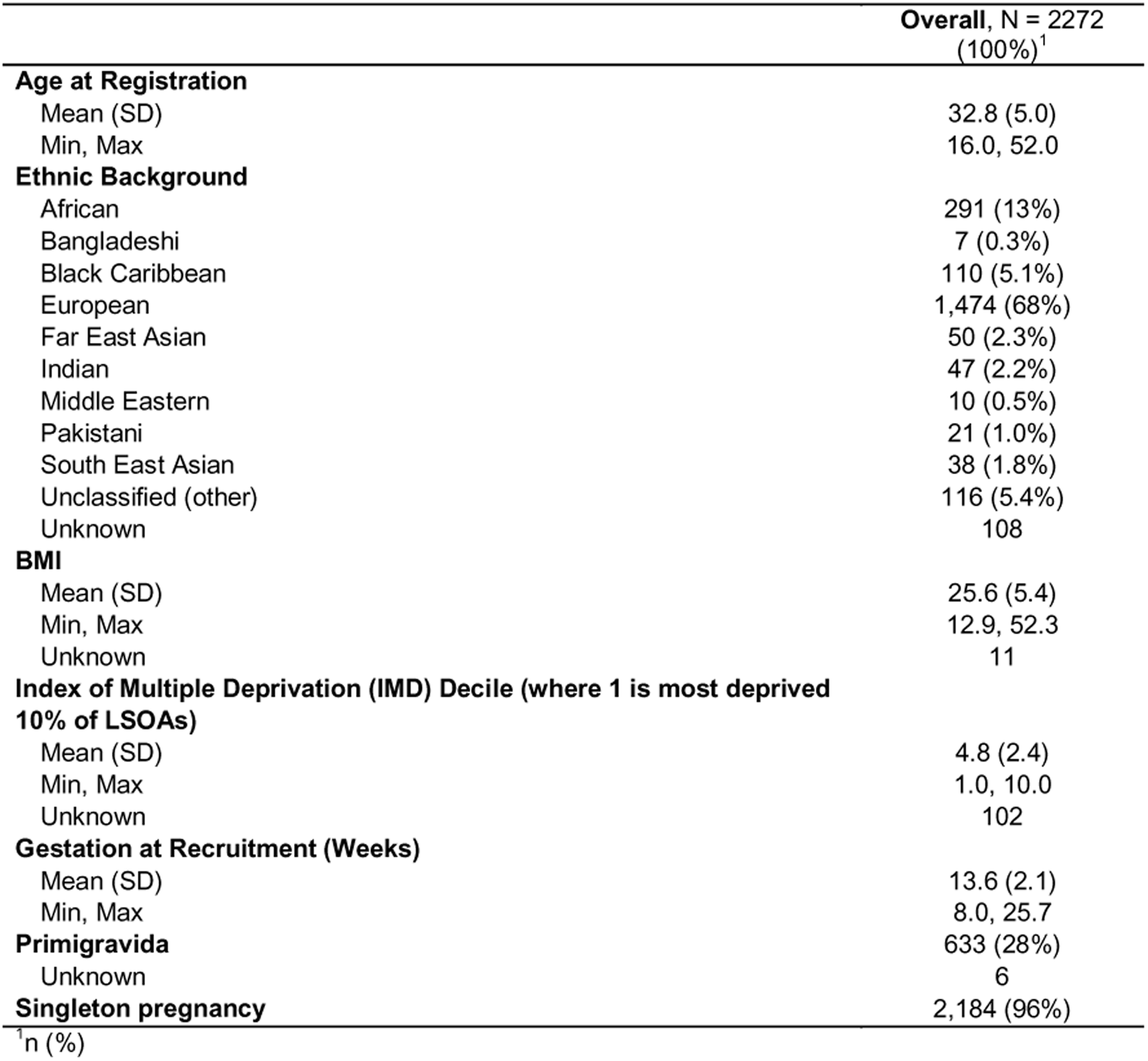
Summary of the sociodemographic and anthropometric characteristics of members of the INSIGHT study cohort.

Additionally, medical history, current medications, and key behavioural and social exposures, were captured both for risk stratification and because of their potential roles in causal pathways to preterm birth. Pre-existing conditions included chronic hypertension, asthma, type 1 and type 2 diabetes, autoimmune disease, chronic renal disease, and chronic viral infection, each of which may independently elevate the risk of adverse pregnancy outcomes or influence clinical management decisions during pregnancy. Medications at enrolment included antihypertensives, steroids, immunosuppressive agents, and antibiotics, etc. and were documented as markers of disease severity and treatment burden. Behavioural exposures (smoking, recreational drug use, and history of domestic violence) and infection-related history (recurrent urinary tract infections and bacterial vaginosis) were recorded because of their well-established links to preterm birth and other adverse perinatal outcomes. A summary of these variables and their completeness is provided in Supplementary Table A. Figure 2 shows, using lower layer super output areas (LSOAs), the general geographic locations of participants indicating the broad geographical reach of the study.

**Figure 2:**
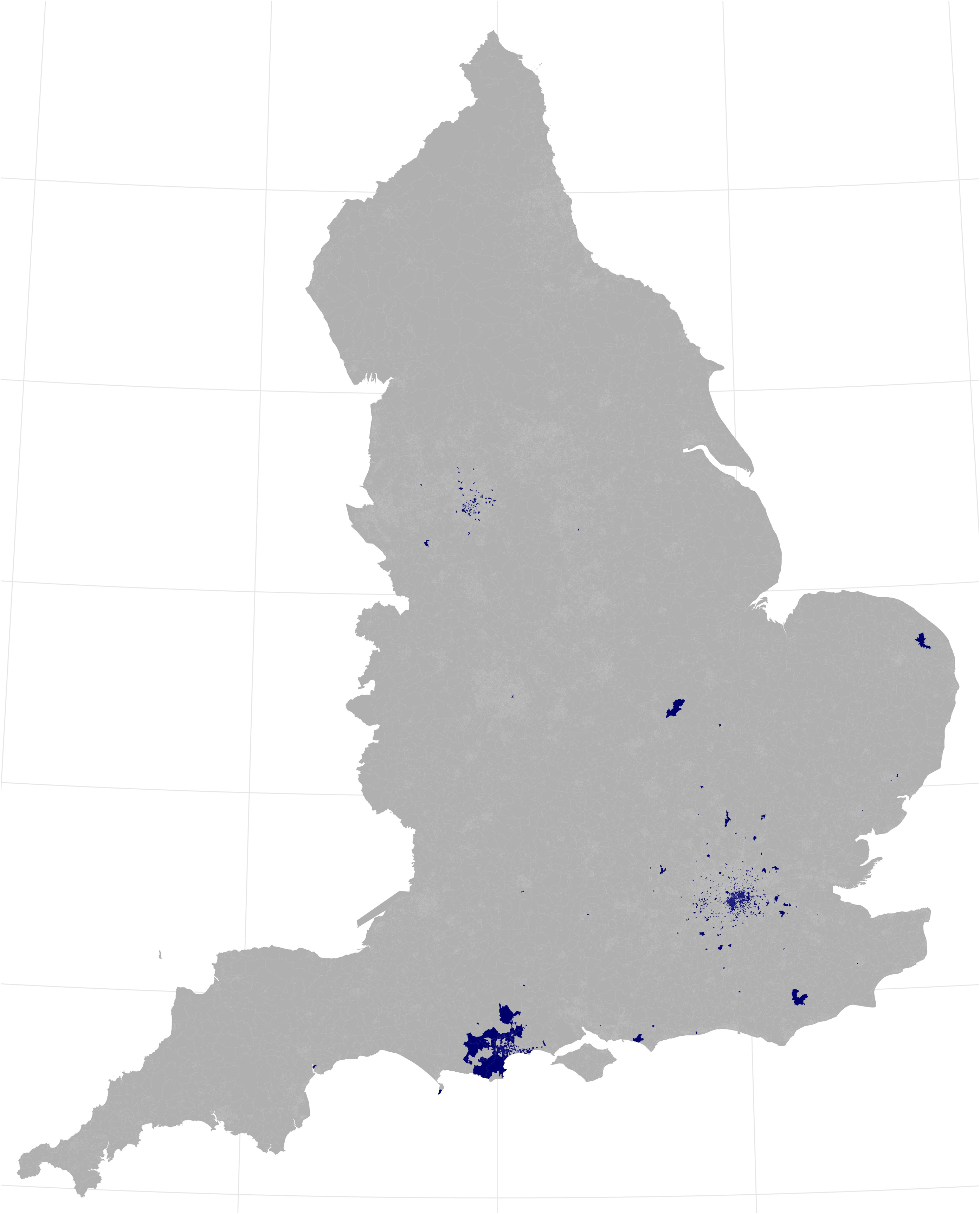
A map indicating (navy blue shading) lower layer super output areas (LSOAs) of participants in the INSIGHT study.

#### b. Risk Factors for Preterm Birth

Within the INSIGHT study, a focused set of established clinical risk factors for spontaneous preterm birth were used. The presence/absence of these factors to designate participants as being at either high or low risk of preterm birth at enrolment. High risk was defined as (i) previous PTB or second-trimester loss between ≥ 16^+0^ and ≤ 37^+6^ weeks’ gestation; (ii) previous preterm prelabour rupture of membranes (PPROM) at ≤ 37^+6^ weeks; (iii) a short cervical length (≤ 25 mm) measured by ultrasound between 18^+0^ and 24^+0^ weeks; (iv) prior cervical procedures (large loop excision, laser conisation, cold knife conisation, trachelectomy, etc.); (v) multiple pregnancy; and (vi) known Müllerian abnormalities.

The frequency of each risk factor in the cohort is summarised and the overlap between risk factors is visualised using an UpSet plot (Figure 3) and frequency of individual risk factors is in Supplementary Table B. Risk factors were not mutually exclusive: cervical-related factors (short cervix and/or prior cervical procedure) often co-occurred with a history of previous preterm birth/mid-trimester loss, and some participants had combined historical and current-pregnancy risk factors. Figure 3 highlights the most common intersections and provides a clear overview of multimorbidity of risk. The most common risk factor was a history of cervical procedures, the least common was a uterine variant. Of the 969 high risk women with a recorded cervical measurement between 18^+0^ and 24^+0^ weeks gestation, only 194 (20%) had a recorded cervical length of ≤ 25 mm.

**Figure 3:**
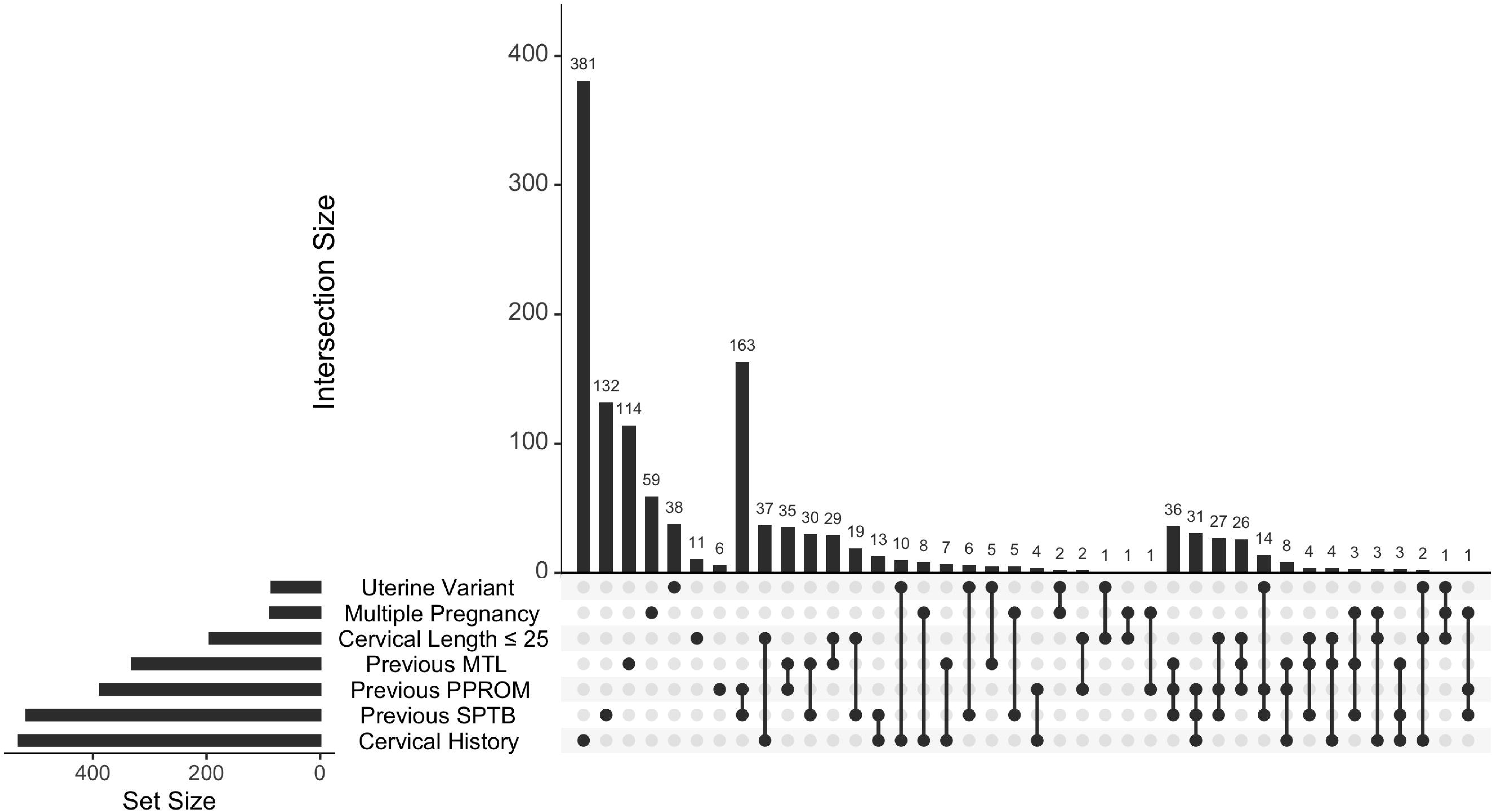
An overview of the presence of the different risk factors used to classify participants as being at high risk of preterm birth. The plot illustrates how often each risk factor occurred irrespective of other co-occurring risk factors (set size) and how often the different risk factors co-occurred (intersection size). Solid black circles indicate which risk factor(s) were present in the group under consideration, black lines connect circles in groups where more than one risk factor was present. For example, 381 participants had cervical history as their only risk factor (first column) while 163 participants had both a previous PPROM and a previous SPTB as risk factors (eight column). Abbreviations: SPTB = Spontaneous Preterm Birth, MTL = Mid-trimester loss, PPROM= preterm premature rupture of the fetal membranes.

#### c. Outcomes

A broad range of data relating to delivery outcomes were captured, with a subset of this summarised in Table 2, stratified by birth type (iatrogenic preterm, spontaneous preterm, and term) to provide a cohort-level overview of general outcomes.

**Table 2:**
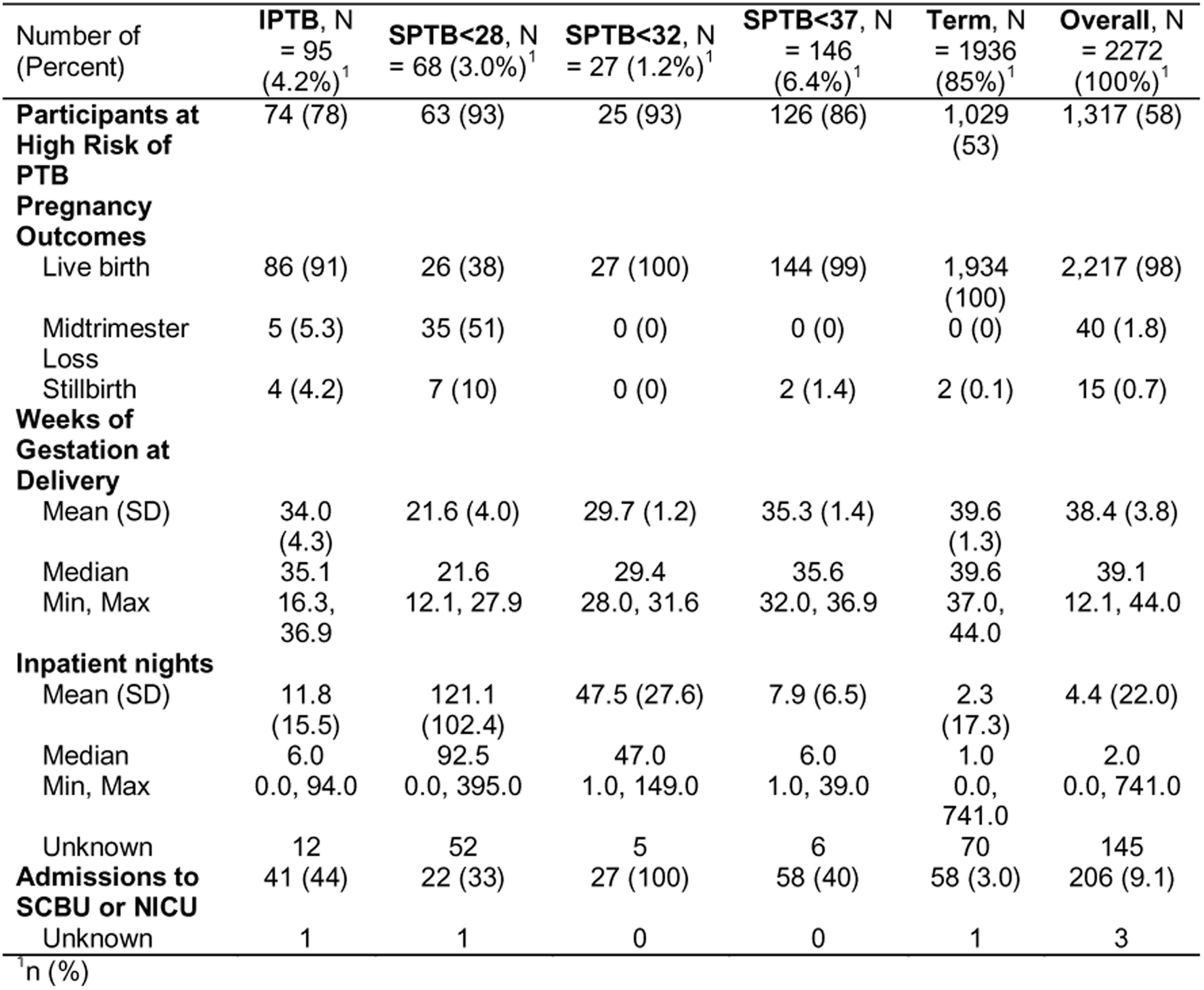
Pregnancy and delivery related outcomes for participants of the INSIGHT study. Outcomes are stratified by birth type: IPTB = iatrogenic preterm birth, SPTB<28 = spontaneous preterm birth before 28 weeks (extremely preterm), SPTB<32 = spontaneous preterm birth before 32 weeks (very preterm), SPTB<37 = spontaneous preterm birth before 37 weeks (moderate preterm), and Term. Definitions are as defined by the World Health Organisation (WHO). Other abbreviations: PTB = preterm birth, SCBU = special care baby unit, NICU = neonatal intensive care unit. The final column in the table, Overall, shows total counts irrespective of birth type.

Beyond the variables presented in Table 2, a detailed set of treatment and delivery-related data were collected. Preventive and acute interventions were recorded, including tocolysis, antenatal corticosteroids, antibiotics, progesterone, magnesium sulphate for neuroprotection, amniocentesis, and in utero transfer, as these reflect both clinical decision-making and the acuity of the presentation. Measures of healthcare utilisation (antenatal day unit attendances; antenatal and postnatal inpatient nights) were included to capture the resource burden associated with threatened and actual preterm birth. Labour and delivery characteristics, onset of labour, augmentation and its recorded indications, timing of membrane rupture, and mode of delivery, were documented to enable detailed characterisation of delivery pathways.

Given the central role of infection and inflammation in the pathophysiology of preterm birth, infection and inflammation markers (bacterial vaginosis diagnosis, elevated C-reactive protein or white cell count, positive midstream urine, high vaginal swab, or blood culture, and placental histological evidence of chorioamnionitis) were also recorded. Maternal outcomes included blood loss, pyrexia, and timing of discharge. Finally, a broad set of neonatal outcomes was collected, birthweight, Apgar scores, major congenital abnormality, infection, respiratory and neurological morbidities, ongoing oxygen requirement or neonatal unit care at 28 days, transfers, and neonatal death, to characterise the short-term morbidity burden for infants across gestational age groups. Together, these variables provide the basis for subsequent analyses examining not only when and how births occurred but also the clinical context, interventions received, and maternal and neonatal outcomes associated with each pathway.

### 8. Sample Collection

Biological samples collected longitudinally across pregnancy included maternal blood and cervicovaginal specimens. Maternal venous blood was processed for serum, plasma, and buffy coat isolation. Cervicovaginal samples were collected during a speculum examination performed by trained clinicians, using a defined clinical procedure (12). Vaginal swabs were obtained from predefined anatomical sites (high vaginal/posterior fornix and low–mid vagina), and endocervical samples were collected using a cytobrush. Swabs were processed for cell pellet and fluid fractions and, where relevant, for microbiological analyses, with samples stored in appropriate transport media, including liquid Amies and TE buffer. Up to six vaginal swabs could be collected at a single visit.

Samples were obtained at recruitment and again around the time of the mid-pregnancy anomaly scan, with additional sampling at antenatal routine or specialist visits, where applicable and consented. We summarize the total number of samples collected across participants, stratified by birth outcome for the participant in Figure 4, additional information on names of samples and storage notes etc. are available in Supplementary Table C.

**Figure 4:**
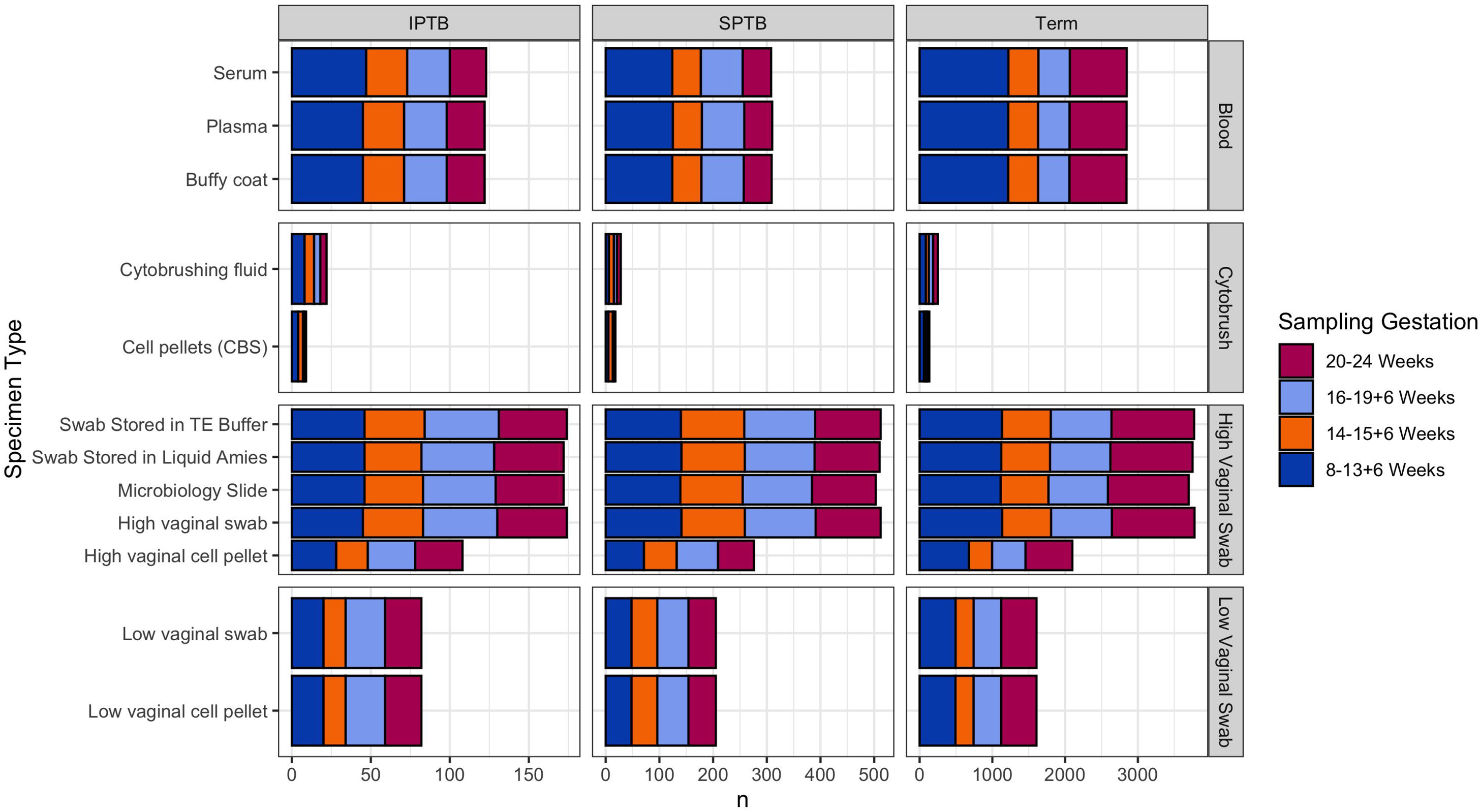
shows the distribution of collected specimen types stratified by primary pregnancy outcome (iatrogenic preterm birth, spontaneous preterm birth, and term birth). Hezelgrave et al. (12) can be referred to for further methodological details on sample collections and protocols. Full notes on names of samples and storage notes etc. are in Supplementary Table C.

## FINDINGS TO DATE

Research outputs generated within the INSIGHT study to date fall into three broad thematic areas.

The first includes outputs which investigate the predictive capabilities of RNA profiling and demonstrating the promise of blood-based, transcriptomic prediction ahead of clinical presentation. Camunas-Soler *et al.* showed that maternal plasma cell-free RNA (cfRNA) can carry a signature that is detectable months before early and very early sPTB (13). In parallel, Rasmussen *et al.* (2022) reported that cfRNA can track pregnancy progression across, placental, maternal and fetal biology and can be used to predict pre-eclampsia (14). Shook and Edlow’s Nature commentary places this body of work in a clinical context and highlights the promise of deploying blood test to predict pregnancy complications (15).

The second thematic area of outputs focuses on the cervicovaginal ecosystem as a determinant of preterm birth biology and risk. Flaviani *et al.* demonstrated that integrating cervicovaginal microbiota and metabolomic profiles sampled in early pregnancy can predict sPTB risk with evidence for interactions between microbes, metabolites, and host immune response (16). Hezelgrave *et al.* quantified cervicovaginal natural antimicrobial expression and found that alterations in innate immune response proteins in early pregnancy are predictive of sPTB (12). This was complemented by work by Chin-Smith *et al.*, who quantified the presence and order of abundance of key cervicovaginal host-defence peptides in early pregnancy and investigated associated regulatory networks (17). Extending the cellular component, Mohd Zaki *et al.* characterised immune cell composition in pregnancy and reported a relationship between cervical neutrophils and specific microbiome species through transcriptomic work (18). Finally, Laleye et al. (19) focussed on the role of the maternal inflammatory proteome during pregnancy in predicting risk of spontaneous preterm birth, highlighting 16 inflammation related proteins as being key predictors of risk.

Finally, a set of outputs have been produced which focusing on connecting microbial metabolism to clinically relevant phenotypes with a focus on real-world prediction of preterm birth. Horrocks *et al.* used nuclear magnetic resonance metabolomics to characterise metabolic strategies and symbioses among bacterial vaginosis (BV) associated bacteria, helping to identify which bacterial interactions may generate metabolites previously linked to BV and preterm birth risk (20). Ridout *et al.* explored the clinical utility of cervical length and quantitative foetal fibronectin of predicting preterm birth risk in high-risk women with cervical cerclage, finding evidence that supported the continued use of these metrics in management decisions for women at risk of treatment (cerclage) failure. (21)

In addition to these direct outputs, the INSIGHT study was linked to the SuPPoRT trial (a multi-centre randomised controlled trial comparing cervical cerclage, cervical pessary and vaginal progesterone, for the prevention of preterm birth in women who develop a short cervix) (22,23) so that INSIGHT participants with a short cervix could be referred into the SuPPoRT trial.

INSIGHT samples and data continue to be analysed for its primary purpose with eight ongoing projects focussed on prediction of preterm birth.

## STRENGTH AND LIMITATIONS

### 1. Strengths

INSIGHT was a large, prospective cohort study conducted over 10 years of recruitment. The design enabled detailed longitudinal phenotyping, providing a strong framework for exploring the biological mechanisms underlying sPTB. By including both high-risk and low-risk pregnant participants, the study captured the full spectrum of risk, facilitating comparative analyses and supporting early biomarker discovery. At its conception, it was one the first studies set up in the UK to consider longitudinal recruitment for preterm birth clinics, a follow on from a smaller proof of principle study (CLIC, REC 06/Q0704/66, (24)). High-risk status was defined using clinically relevant, pre-specified criteria. Additionally, recruitment across multiple NHS sites further enhanced population diversity and strengthened external validity (Figure 2); recruitment at St Thomas’ Hospital particularly reflects the diverse population composition of the local area, Lambeth. Finally, the prospective nature of the study minimised recall bias, while linkage to routinely collected clinical data ensured robust and comprehensive ascertainment of pregnancy and neonatal outcomes.

### 2. Limitations

Recruitment was based in secondary and tertiary care hospital settings, which may limit generalisability to populations managed exclusively in primary care or non-hospital settings. Recruitment was also uneven across sites, with the majority of participants enrolled in London at GSTT, potentially influencing the demographic and clinical profile of the cohort. Identification of high-risk participants through specialist preterm birth clinics may have further enriched the cohort for individuals with more complex or severe risk profiles. As with many longitudinal pregnancy cohorts, attrition over time resulted in incomplete data for some participants, particularly those who transferred care and were lost to follow-up. Lastly, biological sampling was conducted only up to a prespecified gestational age rather than through to birth, limiting assessment of late-pregnancy trajectories; this limitation is being addressed in the ongoing INSIGHT-2 study (25).

### 3. Lessons learned

The creation and maintenance of a long-running pregnancy cohort highlighted the importance of designing study protocols with sufficient flexibility to accommodate evolving scientific priorities, while maintaining clear documentation to support transparency and interpretability. Embedding recruitment within routine clinical care pathways facilitated identification of eligible participants but required sustained engagement with clinical teams to ensure consistency across sites and over time.

Future research builds on this cohort through the INSIGHT-2 study (25), which extends follow-up to include children born from these pregnancies and broadens the scientific focus beyond spontaneous preterm birth to pregnancy complications more generally. This expansion is supported by the collection of additional biological samples, including peripheral blood mononuclear cells (PBMCs) from lithium heparin, cord blood, placental tissue, and child blood for plasma and PBMC isolation, alongside extended sampling time points into late gestation beyond the original pre-24-weeks’ gestation window. INSIGHT-2 also incorporates more comprehensive data collection on lifestyle factors, mental health, dietary intake and infant feeding intentions, laying the foundations for mechanistic and life-course research into maternal and child health outcomes.

Importantly, sustained involvement of patient and public involvement and engagement (PPIE) members was critical to both recruitment and retention, informing patient-facing materials and study procedures to make participation acceptable and feasible. The INSIGHT cohort emphasised the value of involving PPIE contributors from the earliest stages of study conception. For future studies, we suggest establishing an independent steering committee with PPIE representation of the different recruitment groups and external experts to provide oversight, continuity and critical external perspectives as studies evolve. This is in place for the INSIGHT-2 study.

Finally, for all pregnancy cohorts, we recommend explicitly incorporating consent for linkage of maternal and child data to routinely collected health, educational and social care datasets, which enables more comprehensive life-course research while minimising participant burden. This was not originally included in the INSIGHT consent form, although it was introduced for maternal data in 2018 and extended to child data in 2021, and both included in INSIGHT-2

## COLLABORATION

The INSIGHT research team welcomes collaborative proposals from investigators seeking to address research questions aligned with the aims of the INSIGHT study, including those involving data linkage and/or analysis of biological samples, through completion and submission of the INSIGHT Collaboration Request Form (Supplementary Material). Access is granted through an application process in which proposals are reviewed by the INSIGHT Oversight Committee, which assesses scientific merit, feasibility, and potential overlap with existing collaborations. Approved projects are conducted under INSIGHT ethics approval and relevant governance arrangements. Researchers are granted permission to use, but not ownership of, INSIGHT data and samples, and any reuse is subject to the terms of the collaboration agreement and applicable ethical, legal and institutional requirements.

## Supporting information

Supplementary Form

## Data Availability

All data produced in the present work are contained in the manuscript.

## FURTHER DETAILS

## FUNDING DECLARATION

Funding for the INSIGHT cohort was provided from Tommy’s Charity (Reg charity no. 1060508); NIHR Biomedical Research Centre (BRC) based at Guy’s and St Thomas’ National Health Service Foundation Trust, the Rosetrees Trust (no. 298582, M303-CD1), Borne Foundation (no. 1167073), and MRC Capital Investment in Human Tissue Banking and Linked Data in Partnership with Charities Award (MR/R014167/1). NH was funded by a NIHR Doctoral Research Fellowship (DRF-2013-06-171).

## ETHICS

INSIGHT (IRAS: 117589) was reviewed and approved by London City and East Research Ethics Committee (REC: 13/LO/0393). The participants provided written informed consent to participate in this study.

## PATIENT AND PUBLIC INVOLVEMENT STATEMENT

Patients attending the preterm birth surveillance clinic at GSTT have been involved in this research from its earliest stages through an established preterm birth PPIE group, active since 2012. As the clinic is a referral centre, PPIE members are drawn from a wide geographic area and not solely from London. This group has helped shape research priorities, refine research questions, and identify outcomes that reflect patient concerns and preferences. Members have provided input into study design and participant materials, including consideration of participant burden and feasibility. They have also advised on recruitment approaches and will contribute to dissemination planning, including decisions about which findings to share with patient communities and in what formats.

## ACKNOWLEDGMENTS

The authors thank the following for their contributions to the INSIGHT study: Abigail Adeosun, Hira Husain, Soline Caprioli (Clinical Research Assistants) Helen Matthews, Lula Shaban, Omoyele E. Fafowora, Opeoluwa Olusoga (Clinical Trial Associates) Annette Briely (Clinical Trials Manager) Deeba Yunus, Kate Duhig, Maju Chandiramani, Matthew Cauldwell, Raghad Al-Mufti (Doctors) Carla Avena-Zampieri (PhD Student) Jo Girling, Latha Vinayakarao, Melissa Whitwell (PIs) Ellie Cheek, Kirsten White (Research Assistants) Ellie Cheek, Kirsten White (Research Assistants) Lisa Hurley (Research Coordinator) Megan Hall, Agnieszka Glazewska, Alexandra Ridout, Helena Watson, Katy Kuhrt, Lisa Story (Research Fellows) Amy Barker, Angela Chiapparino, Crook Alexandria, Debbie Finucane, Declan Symington, Delphine Strub, Eirini Platsa, Falak Diab, Funso Adegoke, Giorgia Dalla Valle, Hilary Thompson, Holly Lovell, Jennifer Hattan, Jenny Carter, Joelle Pike, Judith Filmer, Lynsey Moorhead, Mairi Alexander, Naomi Carlisle, Rachel White, Ruth Cate, Stephanie Grigsby, Vicky Robinson (Research Midwifes) Leanna Brace (Research Nurse) Caitlin Giles (Research Practitioner) Anna Brockbank, Cally Gill (Research Technicians)

In addition to named contributors the authors additionally thank contributing members from the NIHR Research Delivery Network and from King’s Health Partners.

## CONTRIBUTIONS

The following statement makes use of the Contributor Roles Taxonomy (CRediT). RMT, NLH, and AHS contributed to Conceptualisation of the study. RMT, NLH, AHS, and ECS contributed to Methodology of the study. RMT, NLH, AHS, and NS contributed to Investigation. A full list of individuals and groups who contributed to Investigation is given within the acknowledgements section. RMT, ECS, CV, and RJ contributed to Data Curation. RJ contributed to Visualization. RJ and CV contributed to Writing – original draft. RJ, CV, ECS, NS, AHS, NLH, and RMT contributed to Writing - review and editing. RMT, ECS, and NLH contributed to Project Administration. Chat GPT contributed to Writing – review and editing, specifically it was used when revising this manuscript to assist in improving sentence structure, word choice, and clarity of the text.

## COMPETING INTERESTS STATEMENT

RMT reports a funded collaboration and a consulting fee paid by Mirvie Inc. to King’s College London (KCL) for a collaborative project involving cfRNA measurement in a subset of INSIGHT samples. AHS reports a funded collaboration paid by Pregnolia AG to KCL for a study measuring cervical stiffness using the Pregnolia device in a subset of INSIGHT participants.

## Notes

### Competing Interest Statement

RMT reports a funded collaboration and a consulting fee paid by Mirvie Inc. to Kings College London (KCL) for a collaborative project involving cfRNA measurement in a subset of INSIGHT samples. AHS reports a funded collaboration paid by Pregnolia AG to KCL for a study measuring cervical stiffness using the Pregnolia device in a subset of INSIGHT participants.

### Funding Statement

Funding for the INSIGHT cohort was provided from Tommys Charity (Reg charity no. 1060508), NIHR Biomedical Research Centre (BRC) based at Guys and St Thomas National Health Service Foundation Trust, the Rosetrees Trust (no. 298582, M303-CD1), Borne Foundation (no. 1167073), and MRC Capital Investment in Human Tissue Banking and Linked Data in Partnership with Charities Award (MR/R014167/1). NH was funded by a NIHR Doctoral Research Fellowship (DRF-2013-06-171).

### Author Declarations

INSIGHT (IRAS: 117589) was reviewed and approved by London City and East Research Ethics Committee (REC: 13/LO/0393).

### Summary of Updates

There was an error in the count of participants with a previous preterm birth. This has now been corrected.

