## Supplementary Form for "Cohort Profile: Investigation into Biomarkers to Predict Preterm Birth (INSIGHT) -- a Prospective Pregnancy Cohort Focused on Preterm Birth in the United Kingdom"

**INSIGHT & INSIGHT-2**

**Collaboration Request Form**

### Important Note

Please be mindful that, under INSIGHT and INSIGHT-2 ethics approval, samples and data may be analysed for additional research questions through approved collaboration. However, INSIGHT is not a biobank. You are not applying for ownership of data and samples, but for permission to use them to address research questions that are aligned with the aims of the INSIGHT and/or INSIGHT-2 studies.

### Directions

The lead applicant requesting the collaboration should complete this application. The CI of INSIGHT/INSIGHT-2 must be a co-applicant and other relevant PI’s included on a case-by-case basis. The wider INSIGHT/INSIGHT-2 collaborative group must be acknowledged on publications.

If approved, the requested data and samples will be reserved for use by your study for 12 months, with project activation normally anticipated within 6 months. If after one year, no progress is reported, they will be made available for other researchers to request. Any data generated by your study must be submitted to the INSIGHT bioinformatician for archiving as soon as possible. The data will be embargoed until publication of relevant paper or thesis subject to reasonable time limits.

The data analysed must be reviewed by the INSIGHT Research Team before being submitted for publications.

For academic projects there may be a component of cost recovery and a handling charge for retrieving data and samples to cover the administrative costs (commercial applications will require full cost recovery). Collaborators are responsible for covering these costs and all charges that arise for shipping the samples. The courier used for transferring samples must be approved by the committee.

Please note that if your application is approved, you will most likely need to have a short meeting with the INSIGHT-2 bioinformatician to confirm details of required samples/data.

### 1. Applicants

**Principal applicant:**

| Name: |
| --- |
| Organisation: |
| Email: |
| Telephone: |
| Address: |

**Co-applicants:**

| Name: | Role: | Institution/Organisation: |
| --- | --- | --- |

### 2. Project information

| Title: |  |
| --- | --- |
| Planned start date: |  |
| Planned end date: |  |
| Project category: | ☐ Research  ☐ Audit*  ☐ Service Development  ☐ Service Evaluation  ☐ Quality Improvement  ☐ Implementation Science  ☐ Other  **Please confirm the project has received appropriate clinical governance approval and email the approval to the INSIGHT-2 research office along with this application* |

Does the project have ethics under INSIGHT-2?

☐Yes ☐ No *(– if not, specify ethics details below including REC reference number)*

Does the project require recontacting the selected participants? **☐**Yes **☐** No

### 3. Project timeline

Is there a specific date by which you require the samples/data?

Please be aware that standard processing may take up to 6 weeks depending on the request. If you require a large number of samples to be pulled (>100), processing time may exceed this timeline.

### 4. Collaboration type

Does the proposed project have Commercial Partnerships? **☐**Yes **☐** No

If yes, please provide contact name and details:

Is Intellectual Property (IP) likely to arise from this research? **☐**Yes **☐** No

If yes, please provide further details and the contact details of the IP officer for your Institution:

### 5. Funding

Is funding required to complete the project? **☐**Yes **☐** No *(– please move to section 5)*

| Funding Status | Yes | No | Funder/Amount funded/Date |
| --- | --- | --- | --- |
| Planning to apply for funding |  |  |  |
| Applied for funding |  |  |  |
| Outcome of funding application expected date |  |  |  |
| Funded |  |  |  |

### 6. Justification *(no more than 2 pages)*

Outline the background of the study:

*(If a grant has already been approved for this project, please attach the grant application.)*

Outline the hypothesis of the study:

Outline study aims and the sample size calculation:

Primary objective

Secondary objectives

Sample size calculation

References

### 7. Participant type needed

| Specifics | Gestational age range/Infant age range | Total #Subjects | Total #Controls |
| --- | --- | --- | --- |
| ☐ Normal pregnancy |  |  |  |
| ☐ Pregnancy complications |  |  |  |
| ☐ Fetal |  |  |  |
| ☐ Newborn/Child |  |  |  |

| Maternal Age: | ☐ Any | ☐ Specific ages, list: | |
| --- | --- | --- | --- |
| Fetal Gender: | ☐ Any | ☐ Female Only | ☐ Male Only |
| Pregnancy Outcome: | ☐ Term ☐ Preterm ☐ Stillbirth ☐ Fetal Growth Restriction  ☐ Preeclampsia ☐ N/A ☐ Other: | | |
| Child Health Outcome: | ☐ Healthy ☐ T1D ☐ Other: | | |

### 8. Samples request

| Pregnant women samples | | |
| --- | --- | --- |
| Specimen Type | **Number of aliquots required per participant** | **Minimum volume required for assay** |
| ☐ Serum |  |  |
| ☐ EDTA Plasma |  |  |
| ☐ Whole blood |  |  |
| ☐ Ficoll-diluted plasma |  |  |
| ☐ PBMCs |  |  |
| ☐ EDTA Buffy coat |  |  |
| ☐ Microbiology swab – Liquid Amies |  |  |
| ☐ Microbiology swab – Slide |  |  |
| ☐ Microbiology swab – TE Buffer |  |  |
| ☐ Saliva (hormones) |  |  |
| ☐ Saliva (microbiology) |  |  |
| ☐ Saliva (hormones) |  |  |
| ☐ Cytobrushing fluid |  |  |
| ☐ Cell pellet (CBS) |  |  |
| ☐ High vaginal swab |  |  |
| ☐ Cell pellet (HVS) |  |  |
| ☐ Low vaginal swab |  |  |
| ☐ Cell pellet (LVS) |  |  |
| ☐ Amniotic fluid |  |  |

| Placental Samples | | |
| --- | --- | --- |
| Specimen Type | **Number of aliquots required per participant** | **Minimum volume required per assay** |
| ☐ Snap frozen |  |  |
| ☐ RNA Later |  |  |
| ☐ Paraffin slides |  |  |
| ☐ Cell Suspension |  |  |
| Pictures of placenta(s) – *if taken*: ☐ Yes ☐ No | | |

| Infant Samples | | |
| --- | --- | --- |
| Specimen Type | **Number of aliquots required per participant** | **Minimum volume required per assay** |
| ☐ Cord blood plasma |  |  |
| ☐ Cord blood PBMCs |  |  |
| ☐ Infant blood plasma |  |  |
| ☐ Infant blood PBMCs |  |  |

### 9. Data request

Clinical data

Standard information provided with coded-linked specimens comprises of pregnancy outcome/clinical diagnosis, maternal age, gestational age, infant age and infant gender, if available. Select preference:

☐ Standard information only;

☐ Standard information and additional information indicated below;

☐ No clinical data required.

Additional information from data collection forms may be available upon request. Indicate additional information required:

☐ Stress, mental health and life experiences questionnaires

☐ Dietary recall outcomes

☐ First trimester antenatal visit

☐ Second trimester antenatal visit

☐ Third trimester antenatal visit

☐ Delivery details

Please justify your data selection:

### 10. Agreement

**Signature:**

If you are sending this form by email then you should note that in the absence of this signature, the email of this proposal constitutes your personal certification that the details are correct.

**Date:**

**Name** (*on behalf of applicants)***:**

### For internal use only (INSIGHT-2 Team)

**Allocated application number:**

| Stage | Date | Outcome | Comments |
| --- | --- | --- | --- |
| Received application form |  | Completed ☐ Yes ☐ No |  |
| Discussed with Team |  | ☐ Declined  ☐ Declined but can resubmit  ☐ Provisionally accepted  ☐ Accepted |  |
| Re-submitted for review |  |  |  |
| Re-discussed with Team |  | ☐ Declined  ☐ Declined but can resubmit  ☐ Provisionally accepted  ☐ Accepted |  |
| Project agreement requested |  |  |  |
| All contracts/agreements confirmed |  |  |  |
| Data/samples shared |  |  |  |
