## Supplementary Table A for "Cohort Profile: Investigation into Biomarkers to Predict Preterm Birth (INSIGHT) -- a Prospective Pregnancy Cohort Focused on Preterm Birth in the United Kingdom"

| variables | missing_count | missing_percent | Variable_Type |
|---|---|---|---|
| Site | 0 | 0 | Intake |
| Age at Registration | 0 | 0 | Intake |
| Singleton pregnancy | 0 | 0 | Intake |
| Previous spontaneous PTB (<37 weeks) | 0 | 0 | Intake |
| Previous PPROM (<37 weeks) | 0 | 0 | Intake |
| Previous late miscarriage (16-23+6 weeks) | 0 | 0 | Intake |
| Previous cervical surgery (e.g. LLETZ, Cone) | 0 | 0 | Intake |
| Cervical length <25mm this pregnancy | 0 | 0 | Intake |
| Low risk at enrolment | 0 | 0 | Intake |
| Pre-existing hypertension | 3 | 0 132042254 | Intake |
| Asthma | 3 | 0 132042254 | Intake |
| Type 1 diabetes | 3 | 0 132042254 | Intake |
| Type 2 diabetes | 3 | 0 132042254 | Intake |
| Autoimmune disease | 3 | 0 132042254 | Intake |
| Chronic renal disease | 3 | 0 132042254 | Intake |
| Chronic viral infection | 3 | 0 132042254 | Intake |
| Medical history - Other | 3 | 0 132042254 | Intake |
| Antihypertensives | 3 | 0 132042254 | Intake |
| Steroids | 3 | 0 132042254 | Intake |
| Immunosuppressive agents | 3 | 0 132042254 | Intake |
| Antibiotics | 3 | 0 132042254 | Intake |
| Current medications (at time of enrolment) - Other | 3 | 0 132042254 | Intake |
| Height | 9 | 0 396126761 | Intake |
| Weight | 10 | 0 440140845 | Intake |
| Estimated Delivery Date | 0 | 0 | Intake |
| Maternal Blood Group | 789 | 34 72711268 | Intake |
| Ethnic Background | 108 | 4 753521127 | Intake |
| Antiphospholipid Syndrome confirmed positive | 7 | 0 308098592 | Intake |
| Lupus Antibodies confirmed positive | 7 | 0 308098592 | Intake |
| History of 2 or more, proven, recurrent UTIs in pregnancy | 7 | 0 308098592 | Intake |
| Past or present history of GBS | 7 | 0 308098592 | Intake |
| Past or present history of BV | 7 | 0 308098592 | Intake |
| Past or present history of Domestic Violence | 444 | 19 54225352 | Intake |
| Past or present history of recreational drug use | 7 | 0 308098592 | Intake |
| Smoking | 7 | 0 308098592 | Intake |
| Lower super output area | 101 | 4 445422535 | Intake |
| Index of Multiple Deprivation (IMD) Decile (where 1 is most c | 102 | 4 48943662 | Intake |
| Primigravida | 6 | 0 264084507 | Intake |
| Gestation at Recruitment | 0 | 0 | Intake |
| Uterine abnormality | 0 | 0 | Intake |
| Received tocolysis | 17 | 0 67114094 | Outcome |

| | | | |
|---|---|---|---|
| Received steroids for fetal lung maturation | 22 | 0 868535334 | Outcome |
| Received antibiotics | 24 | 0 947493091 | Outcome |
| Received progesterone | 87 | 3 434662456 | Outcome |
| Amniocentesis | 21 | 0 829056455 | Outcome |
| Diagnosed with BV | 21 | 0 829056455 | Outcome |
| Received in utero transfer | 85 | 3 355704698 | Outcome |
| Total no. of Antenatal Day Unit attendances | 26 | 1 026450849 | Outcome |
| Total no. of Antenatal inpatient nights | 25 | 0 98697197 | Outcome |
| Onset of labour | 2 | 0 078957758 | Outcome |
| Labour augmented | 6 | 0 236873273 | Outcome |
| Justification for Labour Augmentation:Preeclampsia | 0 | 0 | Outcome |
| Justification for Labour Augmentation:Obstetric Cholestatis | 0 | 0 | Outcome |
| Justification for Labour Augmentation:Pre-existing/Gestatior | 0 | 0 | Outcome |
| Justification for Labour Augmentation:Antepartum Haemorrl | 0 | 0 | Outcome |
| Justification for Labour Augmentation:Maternal Infection | 0 | 0 | Outcome |
| Justification for Labour Augmentation:Other Maternal Medic | 0 | 0 | Outcome |
| Justification for Labour Augmentation:Suspected fetal growt | 0 | 0 | Outcome |
| Justification for Labour Augmentation:Other suspected fetal | 0 | 0 | Outcome |
| Justification for Labour Augmentation:Post dates | 0 | 0 | Outcome |
| Justification for Labour Augmentation:Malpresentation | 0 | 0 | Outcome |
| Justification for Labour Augmentation:Pre-labour ruptured m | 0 | 0 | Outcome |
| Justification for Labour Augmentation:Other reason | 0 | 0 | Outcome |
| Received magnesium sulphate for fetal neuroprotection | 414 | 16 34425582 | Outcome |
| Antibiotics in labour | 8 | 0 31583103 | Outcome |
| Maternal pyrexia | 15 | 0 592183182 | Outcome |
| Elevated CRP | 19 | 0 750098697 | Outcome |
| Elevated WCC | 19 | 0 750098697 | Outcome |
| Positive MSU | 18 | 0 710619818 | Outcome |
| Positive HVS | 18 | 0 710619818 | Outcome |
| Positive Blood Culture | 17 | 0 67114094 | Outcome |
| Placenta histology shows Chorioamnionitis | 17 | 0 67114094 | Outcome |
| Blood loss | 97 | 3 829451244 | Outcome |
| Date of discharge | 89 | 3 513620213 | Outcome |
| Total inpatient postnatal nights | 241 | 9 514409791 | Outcome |
| Baby number | 0 | 0 | Outcome |
| Date of delivery | 0 | 0 | Outcome |
| Time of delivery | 11 | 0 434267667 | Outcome |
| Gestation at delivery (w) | 0 | 0 | Outcome |
| Gestation at delivery (d) | 0 | 0 | Outcome |
| Date of ruptured membranes | 18 | 0 710619818 | Outcome |
| Time of ruptured membranes | 27 | 1 065929728 | Outcome |
| Mode of delivery | 0 | 0 | Outcome |

| | | | |
|---|---|---|---|
| Pregnancy Outcome | 0 | 0 | Outcome |
| Gender | 28 | 1 105408606 | Outcome |
| Birthweight | 13 | 0 513225424 | Outcome |
| Apgar Score 1 min | 188 | 7 422029214 | Outcome |
| Apgar Score 5 min | 189 | 7 461508093 | Outcome |
| Major congenital abnormality | 7 | 0 276352152 | Outcome |
| Admission to SCBU or NICU | 6 | 0 236873273 | Outcome |
| Respiratory Distress Syndrome | 0 | 0 | Outcome |
| Thrombocytopenia | 0 | 0 | Outcome |
| Pulmonary Hypertension of the Newborn | 0 | 0 | Outcome |
| Necrotising Enterocolitis | 0 | 0 | Outcome |
| Meconium Aspiration Syndrome | 0 | 0 | Outcome |
| Pneumothorax | 0 | 0 | Outcome |
| Retinopathy of Prematurity | 0 | 0 | Outcome |
| Hypoxic Ischemic Encephalopathy | 0 | 0 | Outcome |
| Intraventricular Haemorrhage | 0 | 0 | Outcome |
| USS abnormality of brain | 0 | 0 | Outcome |
| Oxygen at 28 days | 0 | 0 | Outcome |
| Neonatal Unit at 28 days | 0 | 0 | Outcome |
| Positive Kleihauer | 0 | 0 | Outcome |
| Neonatal death | 0 | 0 | Outcome |
| Positive culture of infection in first 48 hours | 8 | 0 31583103 | Outcome |
| Date of discharge from hospital | 155 | 6 119226214 | Outcome |
| Number of inpatient nights | 155 | 6 119226214 | Outcome |
| Pregnancy Outcome Status | 0 | 0 | Outcome |
| Spontaneous onset of labour resulting in delivery <37/40 | 1 | 0 039478879 | Outcome |
| Spontaneous onset of labour resulting in delivery <34/40 | 0 | 0 | Outcome |
| Threatened Preterm Labour | 2 | 0 078957758 | Outcome |
| Premature Prelabour Rupture of Membranes | 1 | 0 039478879 | Outcome |
| Preeclampsia | 4 | 0 157915515 | Outcome |
| Obstetric Cholestasis | 4 | 0 157915515 | Outcome |
| Gestational Diabetes | 4 | 0 157915515 | Outcome |
| Antepartum Haemorrhage | 4 | 0 157915515 | Outcome |
| Maternal Death | 4 | 0 157915515 | Outcome |
| Pregnancy Complications Other | 4 | 0 157915515 | Outcome |
