## Supplementary Tables B and C for "Cohort Profile: Investigation into Biomarkers to Predict Preterm Birth (INSIGHT) -- a Prospective Pregnancy Cohort Focused on Preterm Birth in the United Kingdom"

| **Characteristic** | **N = 2,272**^1^ |
| --- | --- |
| Previous SPTB | 517 |
| Previous MTL | 331 |
| Previous PPROM | 387 |
| Cervical Length ≤ 25 |  |
| No | 1,566 |
| Yes | 194 |
| Not Measured | 512 |
| Cervical History | 530 |
| Multiple Pregnancy | 88 |
| Uterine Variant | 85 |
| High Risk | 1,317 |
| ^1^n | |

***Supplementary Table B:*** *Cohort risk type frequency. Abbreviations: SPTB = Spontaneous Preterm Birth, MTL = Mid-trimester loss, PPROM= preterm premature rupture of the fetal membranes.*

| **Source** | **Sample** | **Name for specimen** |
| --- | --- | --- |
| Blood in serum separator tube (SST) | Serum | Serum |
| Blood in tube containing ethylenediaminetetraacetic acid (EDTA) | Plasma | EDTA plasma |
|  | Buffy coat | EDTA buffy coat |
| Microbiology swab 1 | Slide | Microbiology slide from high vaginal swab |
|  | Liquid Amies | High vaginal Amies (charcoal) medium swab |
| Microbiology swab 2 | TE buffer | High vaginal swab in TE buffer |
| High vaginal swab | High CVF (fluid) | High cervicovaginal fluid |
|  | High cell pellet | Cell pellet from high vaginal swab |
| Low vaginal swab | Low CVF (fluid) | Low cervicovaginal fluid |
|  | Low cell pellet | Cell pellet from low vaginal swab |
| Cytobrush | Cytobrushing fluid | Fluid from endocervical cytobrush |
|  | Cell pellet CBS | Cell pellet from endocervical cytobrush |

***Supplementary Table C:*** *Extended description of collected sample types and obtained specimens for long-term storage as part of the INIGHT study.*
